# Task-Induced Mental Fatigue and Motivation Influence Listening Effort as Measured by the Pupil Dilation in a Speech-in-Noise Task

**DOI:** 10.1101/2022.01.04.22268734

**Authors:** Defne Alfandari Menase, Michael Richter, Dorothea Wendt, Lorenz Fiedler, Graham Naylor

## Abstract

**Objectives:** Listening effort and fatigue are common complaints among individuals with hearing impairment (HI); however, the underlying mechanisms, and relationships between listening effort and fatigue are not well understood. Recent quantitative research suggests that the peak pupil dilation (PPD), which is commonly measured concurrent to the performance of a speech-in-noise task as an index of listening effort, may be informative of daily-life fatigue, but it remains unknown whether the same is true for task-induce fatigue. As fatigue effects are known to manifest differently depending on motivation, the main aim of the present study was to experimentally investigate the interactive effects of task-induced fatigue and motivation on the PPD.

**Design:** In a pre-/post-fatigue within-subject design, 18 participants with normal hearing (NH) engaged in a 98-trial-long speech-in-noise task (the ‘load sequence’, approximately 40 min. long), which either excluded or included additional memory demands (light vs. heavy load sequence). Before and after the load sequence, baseline pupil diameter (BPD) and PPD were measured during shorter ‘probe’ blocks of speech-in-noise tasks. In these probe blocks, if participants correctly repeated more than 60% of the keywords, they could win vouchers of either 20 or 160 Danish krones worth (low incentive vs. high incentive). After each probe block, participants reported their invested effort, tendency for quitting, and perceived performance.

**Results:** The BPD in anticipation of listening declined from pre-to post-load sequence, suggesting an overall decrease in arousal, but the decline did not scale with the magnitude of the load sequence, nor with the amount of monetary incentive. Overall, there was larger pre-to post-load sequence decline in PPD when the load sequence was heavy and when the monetary incentives were low. Post-hoc analyses showed that the decline in PPD was only significant in the heavy-load sequence-low reward condition. The speech-in-noise task performance, self-reported effort, and self-reported tendency to quit listening did not change with the experimental conditions.

**Conclusions:** This is the first study to investigate the influence of task-induced fatigue on BPD and PPD. Whereas BPD was not sensitive to the magnitude of previous load sequence and monetary incentives, the decline in PPD from pre-to post-load sequence was significant after the heavy load sequence when the offered monetary incentives were low. This result supports the understanding that fatigue and motivation interactively influence listening effort.

---

Perceiving speech in noisy environments is known to be more effortful for individuals with hearing impairment (HI) than those with normal hearing (NH; e.g. Dwyer, Firszt, & Reeder, 2014). Such daily listening effort, which taxes cognitive and physiological resources (Pichora-Fuller et al., 2016), is assumed to give rise to listening-related fatigue (Holman, Drummond, Hughes, & Naylor, 2019). School children with HI are known to report more fatigue as compared to their peers (Bess & Hornsby, W., Y., 2014). In adults with HI, daily effort is related to increased need for recovery and sick leave from work (Kramer, Kapteyn, & Houtgast, 2006; Nachtegaal et al., 2009). Despite frequent reports of effort and fatigue among individuals with HI, the relationships between and mechanisms underlying listening effort and fatigue are not well understood (Hornsby, Naylor, & Bess, 2016; Hornsby & Kipp, 2016; Pichora-Fuller et al., 2016; Schneider, Bernarding, Francis, Hornsby, & Strauss, 2019).

Listening effort has widely been assessed using pupillometry (Kramer, Kapteyn, Festen, & Kuik, 1997; Naylor, Koelewijn, Zekveld, & Kramer, 2018; Zekveld, Koelewijn, & Kramer, 2018). The recording of the pupil size is commonly combined with a speech-in-noise (SIN) test, where participants listen to a sentence in noise and repeat the presented sentence or keywords (Nielsen & Dau, 2011; Portnuff & Bell, 2014; Winn, Wendt, Koelewijn, & Kuchinsky, 2018). In this paradigm the intensity difference between the target sentence and background noise can be varied to estimate the signal-to-noise ratio (SNR) that is required for an individual to correctly perceive a certain percentage of sentences (i.e., the speech reception threshold; SRT). The average pupil diameter during a defined period (e.g., 1 s) before the presentation of the target sentence is referred to as the baseline pupil diameter (BPD), and the maximum pupil size relative to baseline elicited during a trial is referred to as the peak pupil dilation (PPD). Whereas the SIN test is informative of the ability of an individual to perceive speech in noise, pupillometry has been argued to capture the processing load (effort) that it takes an individual to perform the task.

Most research to quantify listening effort with pupillometry has focused on the relationship between listening difficulty and PPD (see Zekveld et al., 2018 for a review). During SIN tests with background babble noise, the PPD has been shown to be largest where speech recognition is around 50 % (SRT50), and lower for both higher and lower SNRs (Ohlenforst et al., 2018; Wendt, Koelewijn, Książek, Kramer, & Lunner, 2018). The increase in PPD with increasing difficulty at the left-hand side of the SRT – PPD inverted U-shaped curve is in line with the well-known relationship between task demands and effort, where effort increases with increasing task demands, as long as the task is within capacity (Brehm & Self, 1989; Richter, Gendolla, & Wright, 2016). The decrease in PPD at the right-hand side of the curve has previously been interpreted as “giving up” by the participants due to the task not being perceived as achievable (Ohlenforst et al., 2018)

In a recent study, the PPD has been shown to be informative of daily-life fatigue (Wang et al., 2018). In that study, a large sample of adults with NH and HI performed a SIN test targeting a 50 % correct response criterion while their pupil diameter was being recorded. A significant negative correlation was found between PPD and self-reported daily-life fatigue. This correlation was independent of hearing acuity. This result suggests that individual differences in daily-life fatigue may be related to individual differences in PPD, but the mechanism driving this association remains unclear. While highly suggestive, the study of Wang et al. (2018) says nothing about how listening effort may interact with task-induced (i.e. transitory) rather than persistent fatigue. Meanwhile, it highlights the potential for confounds arising from daily variations in fatigue, in experiments aiming to study task-induced fatigue.

Mental fatigue is well known to arise after a period of sustained effortful exertion, expressing as a subjective struggle to maintain the exertion required (Boksem, Meijman, & Lorist, 2006; see Müller & Apps, 2019 for a review). Fatigue has previously been associated with decreased arousal and hence decreased BPD in cognitively challenging tasks (Hopstaken, van der Linden, Bakker, & Kompier, 2015a). In previous SIN tasks, BPD has been observed to decline with time-on-task (Alhanbali, Munro, Dawes, Carolan, & Millman, 2020; Ayasse & Wingfield, 2020; Zekveld, Kramer, & Festen, 2010). Fluctuations in BPD have commonly been attributed to the locus coerulus – noradrenergic functional dynamics (Murphy, Robertson, Balsters, & O’connell, 2011). Although changes in BPD have been argued to reflect a variety of states other than fatigue such as learning, habituation, and task (dis)engagement (Ayasse & Wingfield, 2020; Gilzenrat, Nieuwenhuis, Jepma, & Cohen, 2010; McGarrigle, Dawes, Stewart, Kuchinsky, & Munro, 2017) reduced arousal is considered a hallmark of fatigue (McGarrigle et al., 2017; van der Linden, 2011).

In a fatigued state, whether individuals continue to invest effort is known to depend on their willingness to perform the task at hand (Boksem et al., 2006; Boksem & Tops, 2008; Hockey, 1997, 2010). Indeed, a study using pupillometry during a mentally challenging task showed diminishing PPD over time-on-task in a 2-hour long task, but restoration to initial levels after participants were offered rewards for correct performance (Hopstaken, van der Linden, Bakker, & Kompier, 2015b). This result suggests that fatigue and the willingness to perform well (motivation), as manipulated by rewards, together influence the PPD in mentally challenging tasks.

Previous research with pupillometry has shown a positive relationship between rewards and PPD during the SIN paradigm (Koelewijn, Zekveld, Lunner, & Kramer, 2018, although see 2021). PPD was measured in NH adults during a SIN task in quiet and targeting SRT80 and SRT50, corresponding to low, medium and high listening task demands respectively. In blocks of 25 sentences, participants were promised either 5.0 or 0.2 Euro in exchange for correctly repeating 17 (68%) or more of the sentences. When listening demand was low, the size of the monetary incentive had no effect on the PPD. With background noise present, greater incentive led to larger PPD in both task demand conditions. This suggests that particularly when listening demands are high, motivation may have an influence on listening effort.

The aim of the current study was to experimentally investigate the joint influence of fatigue and motivation on the BPD and PPD during a SIN task in a pre-/ post-fatigue paradigm. Pre- and post-fatigue listening effort was measured during SIN tasks in two motivation conditions. Following previous findings of reduced arousal over time on task in SIN tasks (Alhanbali et al., 2020; Ayasse & Wingfield, 2020; Zekveld et al., 2018, 2010), we predicated that BPD would decline from pre-to post-fatigue (Hypothesis 1). As greater fatigue is thought to lead to greater suppression in effort (Müller & Apps, 2019; Schneider et al., 2019), we expected a larger decline in PPD after larger fatigue (Hypothesis 2). Because in a fatigued state the investment of effort is thought to depend on the willingness to exert effort (Müller & Apps, 2019), we expected that any decrease in PPD from pre-to post-fatigue should be larger when motivation is low (Hypothesis 3). Following the thinking that fatigue lowers the willingness for further effort, we expected that the difference in the decline in PPD between the smaller and greater fatigue should be larger when motivation is low (Hypothesis 4).

## Methods

### Participants

Thirty-two adults with normal hearing (i.e. hearing thresholds were less than or equal to 25 dB HL over the frequencies from 0.25 to 4 kHz for both ears) participated. The data of 18 adults (Mean age = 42; *SD* = 16; 7 female) was included in the analyses. Due to excess missing samples (see trial inclusion criteria below), the data of 12 participants were excluded from the analyses. All participants had normal hearing. Participants reported normal or corrected-to normal vision, Danish as native language, and no neurological or psychiatric disorders. Participants were naïve to the fatigue-related aim of the experiment. This was to avoid interoceptive awareness interfering with the natural motivation to perform during the experiment. Later they were debriefed about the purpose of the experiment. The study was approved by the local ethics committee and all participants provided written informed consent.

### Experimental design

Figure 1 illustrates the experimental design. To investigate the interactive influence of fatigue and motivation on listening effort, a within-subject design with two levels of previous mental load and two levels of monetary incentives was used. Listening effort was measured using pupillometry and self-report during the pre- and post-load sequence probes. In this study the load sequence was only used to induce fatigue. The assumptions that the load sequence induces fatigue and that the load sequence with memorization is more effortful were made. No investigations were made on any pupil behavior during the load sequence.

**Figure 1.**
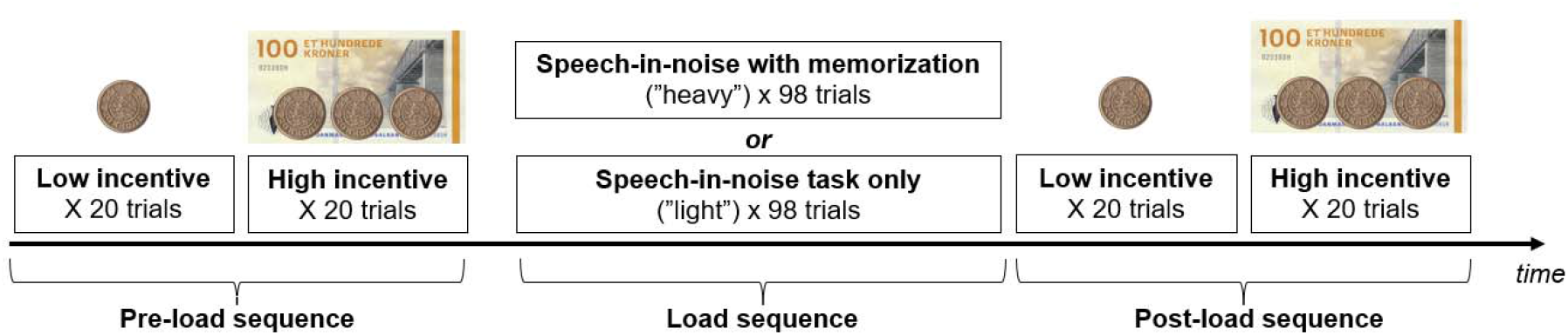
Experimental design. To induce fatigue, a long speech-in-noise test of 98 trials (i.e. “load sequence”) was used. Shorter blocks of 20 trials were administered pre- and post-load sequence to probe the effect of previous mental load on listening effort.

To investigate the influence of previous mental load magnitude on subsequent listening effort, a SIN task with 98 sentences was used (i.e. “load sequence”). The load sequence was approximately 40 minutes in duration. Each participant completed the experiment in 2 sessions. In these sessions, the speech-in-noise task during the load sequence either excluded or included a concurrent memory task (i.e. “light load sequence” and “heavy load sequence”, conditions respectively). In the light load sequence condition, participants only repeated the final words of the sentences that were presented. In the heavy load sequence condition, in addition to repeating the final words of the sentences, participants were also asked to memorize the final words in blocks of 7 sentences. At the end of each 7^th^ sentence, participants were asked to recall the 7 words that were to be memorized.

To probe the influence of previous mental load on subsequent listening effort, both before and after the load sequence, participants performed two SIN tasks (i.e. pre- and post-load sequence ‘probes’). The pre- and post-load sequence probe SIN tasks consisted of 2 blocks of 20 trials each. To investigate the moderating effect of monetary incentives on the influence of previous mental load on subsequent listening effort, in the pre- and post-load sequence blocks, participants were offered either 20 or 160 Danish Krones per block (“high monetary incentive” and “low monetary incentive” conditions, respectively) on the condition that they correctly repeated at least 12 of the 20 sentences (i.e. 60% correct). Per participant, the order of the monetary incentive conditions was the same in the pre- and post-load sequence blocks, and the same across both experimental sessions.

During the load sequence, the SNR within 7-sentence blocks was constant at either +1 or -4 dB, and changed across 7-sentence blocks within sessions in a pre-determined sequence. These SNRs were chosen to span a range of expected recognition rates around 65-100 % (Wendt et al., 2018). To prevent participants from predicting the upcoming SNR during the load sequence and to make the load sequence relatively homogenously taxing across time, the SNR sequence of +1, +1, -4, -4, +1, -4, +1, +1, -4, -4, -4, +1, -4, and +1 dB was created. The SNR was cycled through the sequence with a different starting point for each participant to prevent the possible influence of any sequence-specific effects on subsequent listening effort. To allow comparison of the sessions for the effect of memory load, per participant the SNR order was the same for the light load sequence and heavy load sequence conditions (i.e. in both sessions of the experiment).,

The SNR in the pre- and post-load sequence blocks was individually pre-determined at SRT50 (as described below). This is because, fatigue effects are known to be most evident while performing difficult but achievable tasks (Lagory, Dearen, Tebo, & Wright, 2011). Previous research has shown that the SNR at SRT50 elicited the largest pupil dilation as compared to the other SRT levels, suggesting maximal effort at SRT50 in a rested state (e.g. Wendt, Koelewijn, Książek, Kramer, & Lunner, 2018; Ohlenforst et al., 2018). Therefore, the SNR that corresponds to SRT50 was chosen for the pre- and post-load sequence probe blocks.

### Stimuli and trial sequence

Target stimuli in the pre- and post-load sequence blocks consisted of sentences from the Danish Dagmar-Asta-Tine corpus(DAT; Nielsen, Dau, & Neher, 2014), whereas target stimuli in the load sequence consisted of everyday sentences from the Danish Hearing in Noise Test (HINT; Nielsen & Dau, 2011). The 16-talker babble masker noise consisted of speech segments from 2 female and 2 male talkers. The long-term average frequency spectrum of the masker noise was identical to that of the target speech signal of the corresponding speech corpus, separately for the DAT and HINT material. In this study we only investigated listening effort during the pre- and post-load sequence block. Therefore, only the sequence of trials in the probe blocks are described here.

Figure 2. illustrates the trial sequence in the pre- and post-load sequence blocks. 2.2 seconds after the experimenter pressed a button to start a trial, the babble noise started. The target sentence started 3 seconds after the start of the babble noise. All sentences were 2 seconds long and had the same carrier words “Dagmar tænkte på (∼550 ms) … og en (∼166 ms) … I går (∼333 ms)” (English: Dagmar thought of … and … yesterday”); and 2 keywords (∼450 ms and ∼500 ms, respectively). The babble noise continued for 3 seconds after the ending of the target sentence.

Thereafter participants were asked to repeat the keywords. Answers were counted as correct only if both keywords had been repeated correctly. The experimenter scored the answers by a button-press (“correct or incorrect”), which also triggered the start of the next trial.

**Figure 2.**
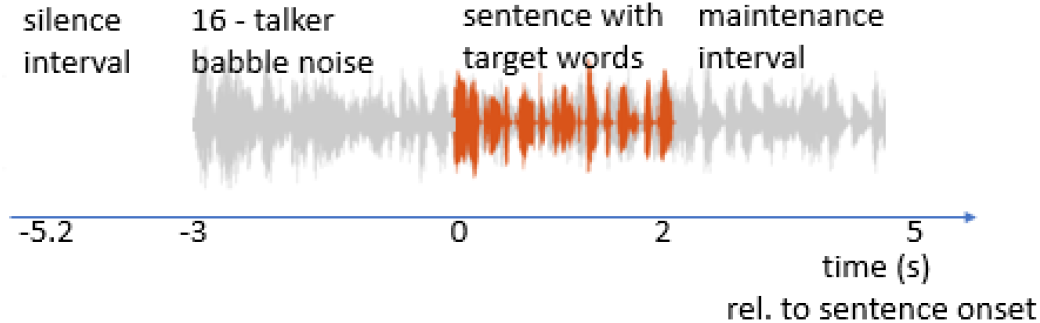
Example trial sequence during the pre-and post-load sequence blocks. All trials started with 2.2 seconds silence, followed by a 16-talker babble noise. Then participants were presented with a sentence (e.g. Dagmar tænkte på en aften o gen festdag I går). The noise continued for 3 seconds after the end of the sentence. Participants were asked to repeat the keywords as accurately and as quickly as possible (e.g. “en aften, festdag”).

### Subjective report scales

After each of the pre- and post- load sequence blocks, participants were asked to report how much effort they mobilized for listening, how well they think they performed, and how much they felt an inclination to quit listening (cf. Koelewijn et al., 2018). Three 100-point visual analog scales were printed in 18 font size on an A4 paper. The questions were (1) “*Hvor meget anstrengte du dig for at høre sætningerne? –(How much effort did you put into hearing the sentences?*”;0 = Ingen, 25 = Lav, 50 = Moderat, 75 = Høj, 100 = Meget høj – EN: 0 = None, 25 = Low, 50 = Moderate, 75 = High, 100 = Very High), (2) “Hvor mange af ordene tror du, at du forstod korrekt?” (*How many of the words do you think you understood correctly*?”; 0 = Ingen, 25 = Mindre end halvdelen, 50 = Halvdelen, 75 = Mere end halvdelen, 100 = Alle – EN: 0 = None, 25 = Less than half, 50 = Half, 75 = More than half, 100 = All), and (3) “*Hvor ofte måtte du opgive at forstå sætningen? (How often did you have to give up understanding the phrase?* (0 = Aldrig, 25 = Mindre end halvdelen af tiden, 50 = Halvdelen af tiden, 75 = Mere end halvdelen af tiden, 100 = Altid – EN: 0 = Never, 25 = Less than half the time, 50 = Half the time, 75 = More than half the time, 100 = all the time). Participants marked their answers on paper.

### Procedure and apparatus

Prior to the experiment participants were informed that the study was about listening effort and motivation. All participants visited the laboratory twice: Once for the session with the light load sequence condition, and once to complete the heavy load sequence condition. The visits were maximum 3 weeks apart. Sessions began at either 9 am or 1 pm, and per participant, both visits were scheduled at the same time of the day to avoid measurements during circadian spikes and to minimize variability of diurnal changes in cortisol (Kirschbaum & Hellhammer, 1989; Lovallo & Buchanan, 2016). Participants were advised to get a good night’s sleep the night before, not take any alcohol 24 hours prior to, and not drink any caffeinated beverages 2 hours prior to the experiment. These precautions were presumed also to minimise any confounding effects from daily-life fatigue as observed by Wang et al. (2018).

All sessions took place in a sound-proof booth. The luminance in the booth was dim (approx. 100 lux) for all participants during all experimental sessions to avoid floor or ceiling effects in the pupil diameter measures. Pupil size was recorded through the Tobii Pro Spectrum eye tracker at a sampling frequency of 1200 Hz. All auditory stimuli were presented via five loudspeakers. The loudspeakers (Genelec 8040A; Genelec Oy, Iisalmi, Finland) were distributed around the participant along an imaginary circle with 1 meter radius. One loudspeaker was positioned in front of the participant (0°) and the four other loudspeakers were positioned at the side and back of the participant (+/-90° and +/-270°). The target stimuli were presented from the loudspeaker in front of the participants, while the 16-talker background noise was presented through the remaining loudspeakers. Pupil size video recording and estimation was performed by a table mounted Tobii Spectrum at 1200 Hz. All presentation of stimuli and collection of data was controlled using software running on MATLAB 2018b (MathWorks, Natick, MA).

First, the speech reception threshold corresponding to 50% correct sentence recognition (SRT50) was determined. This included a practice round and a test round with 20 DAT (Nielsen et al., 2014) sentences each. The background masker was always presented at 70 dB SPL (as averaged across 30 seconds). During the estimation of SRT50 the first sentence was presented at 70 dB and (if the keywords were not repeated correctly) repeated with its level increased by 4dB until the participants repeated all keywords correctly. Sentences 2-4 were presented at levels decreased or increased by 4 dB, depending on whether the participants repeated all the keywords in the previous sentence correctly or not, respectively. The level of the fifth sentence was calculated by taking the average of the levels of the first four sentences and the level at which the fifth sentence would have been presented with a step size of +/-4 dB. For the rest of the sentences, the level of the target sentence was decreased or increased by 2 dB depending on correct or incorrect (incomplete) repetition of the keywords, respectively. The estimated SRT50 was calculated as the average SNR of the last five sentences and what a twenty-first sentence would have been presented at.

Prior to the start of the experiment, to provide participants with an understanding of the listening demands (i.e., SIN at the pre-determined and fixed SRT50), participants performed an additional practice round with 20 sentences in blocks of 10. Participants received feedback from the experimenter per block on how many sentences were repeated correctly.

At the start of each pupillometric data collection session, a 5-point eye-tracker calibration was performed. Thereafter, participants were told that they could earn additional money by correctly repeating minimum 12 of the sentences out of blocks of 20 sentences. Participants were instructed to fixate on a cross on a black screen that was approx. 60 cm away from them and as far as possible to schedule their blinks at the end of their response period. Before each of the 20-trial blocks, participants were informed about the amount of monetary incentives through a verbal message from the experimenter (e.g. “Now we are proceeding with a block of 20 sentences. If you repeat more than 12 correctly, you will receive 160 Danish krones”). In addition, visual reminders of the amount were placed at approximately 20 cm away from the fixation cross on the computer screen. During this session, no feedback was provided to the participants about their performance. Participants were informed beforehand that this session would take approximately 2 hours, but were not informed about how many blocks of trials the experiment had in total. After completing the pre-load sequence probe blocks, participants took a 5-minute break. At the beginning of the load sequence, participants received written and verbal information on the load sequence. To prevent recovery from fatigue, immediately after the load sequence, participants continued with the post-load sequence probe blocks without a break.

### Pre-processing and calculation of the pupil dilation indices

Pupil data were processed using MATLAB R2018b (MathWorks, Natick, MA), similarly to the procedure reported in Wendt et al. (2018). Pupil traces starting 1 s preceding sentence onset until the end of the babble noise were selected for analyses. Traces with more than 30 % of missing data were considered invalid. Per participant, the data from the eye with most valid traces were selected for analysis. The data of a given participant were included in the analyses if 10 or more of the trials per block of 20 trials were valid for all experimental conditions. Pupil diameter values more than 2 standard deviations away from the trial median were coded as blinks. Blinks were later interpolated in a linear fashion. The interpolation started for the samples within 35 ms before and ended for the samples within 100 ms after the blink to account for blink-related changes in pupil size. Later a moving average filter with a window of samples that would fall within 150 ms length was used to smooth the de-blinked trials and to remove any high-frequency artefacts. The trials were averaged per condition, per participant: probe time (pre-vs. post-load sequence) load sequence magnitude (light vs. heavy) x monetary incentive (low vs. high). That is, there were 8 averaged traces per participant. For each averaged trace, the mean value of the trace corresponding to the 1-second interval before the onset of the target sentence was taken as the baseline pupil diameter (BPD). The maximum pupil size during the interval from the onset of the sentence until the end of the presentation of the background babble, minus the BPD, was saved as peak pupil dilation (PPD).

### Statistical analyses

To the best of our knowledge this was the first study to investigate the influence of varying levels of fatigue on the pupil dilation response in varying levels of motivation in a SIN task. Therefore, it was not possible *a priori* to estimate the magnitudes of the main effects of fatigue and its interactions with motivation on the PPD. To capture the contributions of all possible effects to the variance in the PPD, and taking the small sample size into account, we took the approach of testing the mentioned hypotheses with Type III ANOVA’s instead of singling out specific contrasts.

To assess whether there was an overall change from pre-to post-load sequence, t-tests were conducted, where the dependent measures of interest were speech recognition performance, BPD, PPD, self-rated effort, self-rated performance, and self-rated tendency to quit, and the independent variable was probe time (i.e. pre-vs. post-load sequence).

The effects of load sequence magnitude (light and heavy) and monetary incentive amount (low and high) on the change in listening effort from pre-to post-load sequence were investigated by calculating the change in the pupil dilation metrics and self-report scores from the pre-to the post-load sequence blocks (i.e. Δpost-pre).

To control for any effect of the participant’s initial state of fatigue (which may have been different between the two sessions), we calculated the difference scores between pre- and post-load sequence for the pupil and self-report measures. Using these difference scores as dependent variables, we conducted 2-by-2 repeated-measures ANOVAs with the factors load sequence magnitude (light and heavy) and monetary incentive amount (high and low).

## Results

### Speech reception thresholds used in the experiment

The average speech level corresponding to the SRT50 of the participants in a background noise of 70 dB was 65.2 dB (range: 62.6 - 68.4; *SD* = 1.5 dB), i.e. SRT50 = -4.8 dB SNR.

### Performance in the load sequence

For the light load sequence condition, average performance for repeating the last words of the sentences was 83.72 % (range: 67 – 94, *SD* = 7.95), whereas for the heavy sequence, average performance for repeating the last words of the sentences was 79.78 % (range: 66 – 95, *SD* = 9.09). There was no evidence for a difference in repetition performance between the conditions [*t*_*paired*_ (17) = 1.967, *p* = .066]. These results are reported in more detail in Książek et al. (under review).

### Speech recognition performance

Figure 3 shows pre- and post-load sequence speech recognition performance as a function of load sequence magnitude and monetary incentive amounts. The average performance was 51.7 % (*Median* = 52.5, *SD* = 10.9) and did not change from pre-to post-load sequence, [*t*_*paired*_ (17) = 0.695, *p* = .497]. The repeated measures ANOVA with Δpost-/pre-performance score as the dependent variable, and load sequence magnitude (light vs. heavy) and monetary incentive amount (high vs. low) as independent variables showed no main effect of load sequence magnitude, [*F* (1,17) = 0.716, *p* = .409, η_*p*_ ^*2*^ = .040], no main effect of monetary incentive amount, [*F* (1,17) = 0.250, *p* = .624, η_*p*_ ^*2*^ = .014], and no load sequence magnitude x monetary incentive amount interaction effect, [*F* (1,17) < 0.001, *p* = 1, η_*p*_ ^*2*^ < .001]. In sum, there was no significant difference in performance depending on any experimental condition.

**Figure 3.**
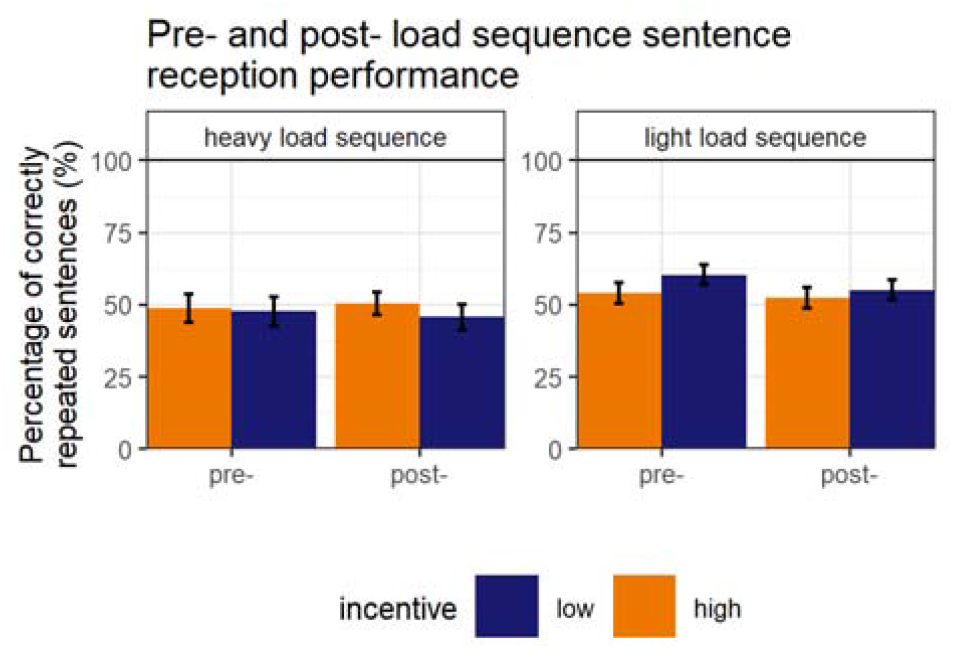
Sentence recognition performance during the pre- and post-load sequence probe blocks as a function of load sequence magnitude and monetary incentive amounts.

### Pupil traces, baseline pupil diameter (BPD), and peak pupil dilation (PPD)

Figure 4A shows the baseline-corrected pre- and post-load sequence pupil traces starting from the beginning of the baseline interval, until the end of the maintenance interval, averaged across the participants, and grouped together according to load sequence magnitude and monetary incentive amount conditions. By visual inspection alone we observe that all traces show a peak around 1 second after the end of the target sentence (i.e., 3 seconds relative to target onset). Furthermore, the peak is larger in the pre-load sequence condition as compared to post-load sequence. The difference between pre- and post-load sequence peaks seems larger for the heavy load sequence condition as compared to that of light, and larger for the low incentive condition as compared to that of high.

**Figure 4.**
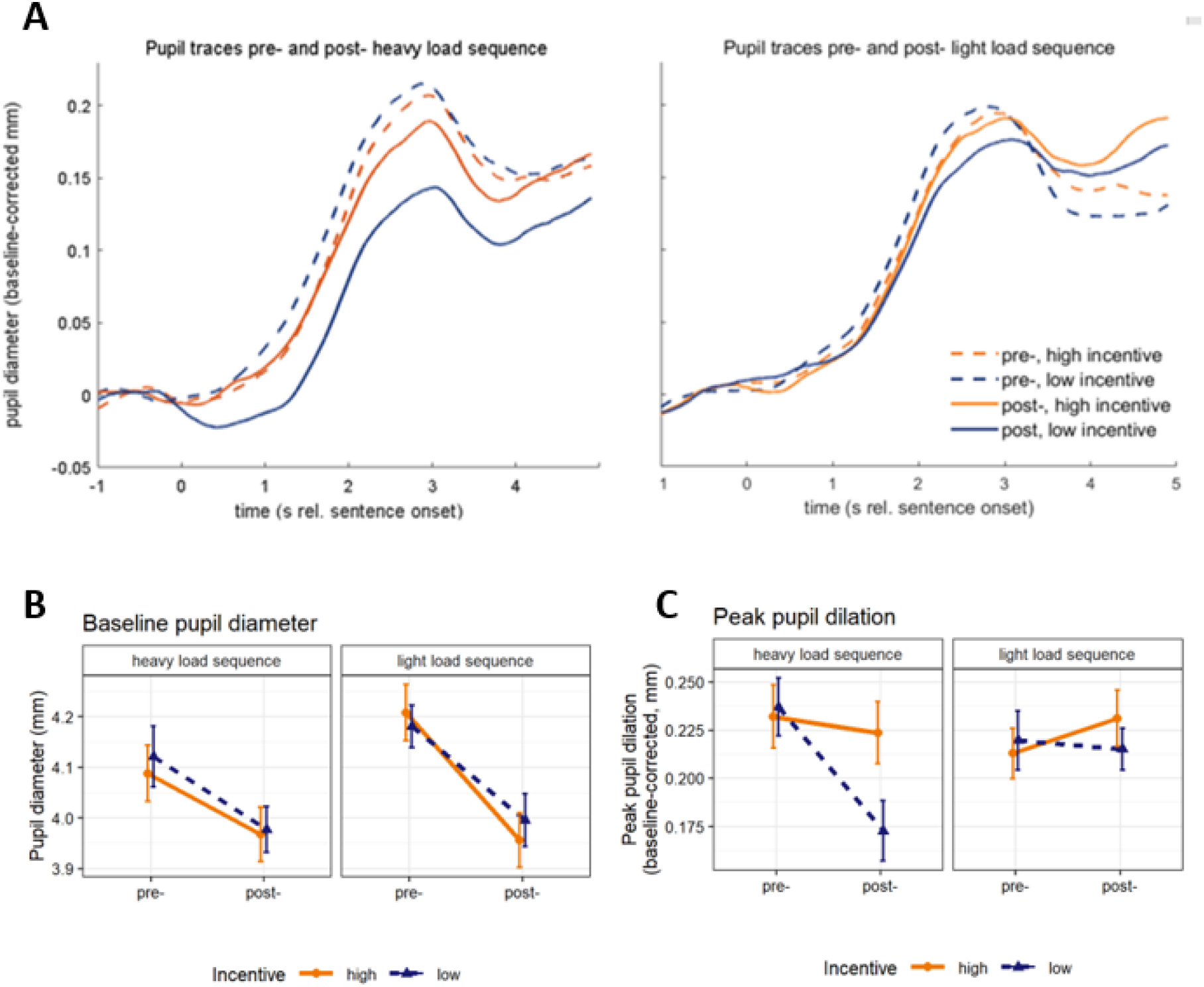
Pre- and post-load sequence (A) baseline-corrected pupil traces (B) the baseline pupil diameter, and (C) baseline-corrected peak pupil dilation as a function of load sequence magnitude and monetary incentive amount. The error bars in (B) and (C) show the within-subject standard error of the mean.

Figure 4.B. shows the pre- and post-load sequence BPD as a function of load sequence magnitude and monetary incentive amount. On average pre-load sequence BPD was larger than post-load sequence BPD, [*t*_*paired*_ (17) = 4.599, *p* < .001]. The repeated measures ANOVA with Δpost-/pre-of BPD as dependent variable, and load sequence magnitude and monetary incentive amount as the independent variables showed no effect of monetary incentive, [*F* (1,17) = 0.639, *p* = .435, η_*p*_^*2*^ < .036], load sequence, [*F* (1,17) = 2.187, *p* = .157, η_*p*_^*2*^ < .114], or monetary incentive x load sequence interaction effect, [*F* (1,17) = 1.2, *p* = .289, η_*p*_^*2*^ < .066].

Figure 4.C. shows the pre- and post-load sequence PPD as a function of load sequence magnitude and monetary incentive amount. Averaged across the monetary incentive and load sequence conditions, overall, the difference in PPD between the pre- and post-load sequence conditions was not significant, [t_*paired*_ (17) = 1.195, *p* = .248]. The repeated measures ANOVA with Δpost-/pre-of PPD as dependent variable, and monetary incentive and load sequence as the independent variables showed a main effect of load sequence, [*F* (1,17) = 4.899, *p* = .041, η_*p*_^*2*^ < .224], as the decline in PPD from pre-to post-load sequence probe was larger after the heavy load sequence as compared to the light load sequence. In addition, there was a main effect of monetary incentive, [*F* (1,17) = 11.886, *p* = .003, η_*p*_^*2*^ < .411], as the decline from pre-to post-was larger when monetary incentives were low. Although the mean difference between the monetary incentive conditions in the decline from pre-to post load sequence was larger after the heavy load sequence as compared to the light load sequence, the monetary incentive x load sequence interaction effect on the Δpost-/pre-of PPD was not significant, [*F* (1,17) = 1.286, *p* = .273, η_*p*_^*2*^ < .070]. However, post-hoc analyses with Tuckey’s method showed that the Δpost-/pre-differed from zero only in the heavy-load-sequence-low-monetary-incentive condition [*p* = 0.05; all other *p’s* > 0.05].

### Subjective ratings of effort, performance, and quitting

Table 1 shows pre- and post-load sequence self-rated effort, performance, and tendency to quit listening as a function of load sequence magnitude and monetary incentive amount. Shapiro-Wilk tests of normality confirmed that all the rating scores were normally distributed [all *p*’s > 0.05].

**Table 1.** Self-rated effort, performance, and tendency to quit listening. Mean (s.d.) across participants

| **Table 1.** |  |  |  |  |  |
| --- | --- | --- | --- | --- | --- |
|  |  | Fatigue manipulation |  |  |  |
|  |  | heavy load sequence |  | light load sequence |  |
| Rating scale<br>(range 0-100) | Probe time<br>Incentives | Pre- | Post- | Pre- | Post- |
| Effort | High | 80 (3.11) | 78.06 (3.34) | 74.72 (3.39) | 75.00 (3.84) |
|  | Low | 79.44 (3.08) | 78.33 (3.11) | 74.44 (3.64) | 75.83 (2.84) |
| Performance | High | 52.50 (4.70) | 49.16 (3.98) | 53.61 (3.63) | 52.55 (4.52) |
|  | Low | 52.22 (4.61) | 51.38 (4.86) | 60.55 (3.78) | 47.50 (4.04) |
| Tendency to quit listening | High | 27.22 (4.85) | 28.06 (3.73) | 25.83 (4.66) | 29.44 (5.21) |
|  | Low | 28.61 (3.81) | 28.33 (4.53) | 23.61 (5.06) | 31.67 (5.53) |

### Self-rated effort

On average there was no evidence for a change in self-rated effort from pre-to post-load sequence, [t_paired_(17) = 0.200; p =.844]. The repeated measures ANOVA with Δpost-/pre- of self-rated effort as dependent variable, and monetary incentive and load sequence as the independent variables showed no effect of load sequence, [*F* (1,17) = 1.253, *p* = .280, η_*p*_^*2*^ = .073], or monetary incentive [*F* (1,17) = 1.916, *p* = .185, η_*p*_^*2*^ = .107]. Nor was there a load sequence x monetary incentive interaction effect, [*F* (1,17) = 0.427, *p* = .523, η_*p*_^*2*^ = .026].

### Self-rated performance

On average there was no evidence for a change in self-rated performance from pre- to post-fatigue, [t_paired_(17) = 1.797; p =.090]. The repeated measures ANOVA with Δpost-/pre- of self-rated performance as dependent variable, and load sequence and monetary incentive as the independent variables showed no effect of load sequence, [*F* (1,17) = 0.773, *p* = .392, η*p 2* = .073], or monetary incentive [*F* (1,17) = 1.909, *p* = .185, η _*p*_ ^*2*^ = .101]. Nor was the load sequence x monetary incentive interaction effect significant, [*F* (1,17) = 4.360, *p* = .052, η_*p*_ ^*2*^ = .204].

### Self-rated tendency to quit listening

On average there was no evidence for a change in self-reported tendency for quitting from pre- to post-fatigue, [t_paired_(17) = 0.200; p =.844]. The repeated measures ANOVA with Δpost-/pre- of self-rated tendency for quitting as dependent variable, and monetary incentive and load sequence as the independent variables showed no effect of load sequence, [*F* (1,17) = 0.961, *p* = .341, η_*p*_^*2*^ = .053], monetary incentive [*F* (1,17) = 1.156, *p* = .697, η_*p*_^*2*^ = .009], or a load sequence x monetary incentive interaction effect, [*F* (1,17) = 0.836, *p* = .373, η_*p2*_ = .047].

## Discussion

The main aim of the current study was to investigate fatigue and motivation effects in pupillometric measures of listening effort. This was realized through the assessment of listening effort in a pre-/post-fatigue experiment, where two levels of load sequence (light and heavy) were used to induce mental fatigue, and applying two levels of monetary incentives (high and low). We hypothesized a decline in BPD from pre- to post-load sequence (Hypothesis 1). Furthermore, following the view that the willingness to invest effort when in a fatigued state depends on the perceived value of investing effort, we hypothesized that any decline in PPD should be larger after the heavier load sequence (Hypothesis 2), and larger with smaller incentives (Hypothesis 3). The difference in decline between the heavy and light conditions should be particularly larger when incentives are low (Hypothesis 4).

Supporting Hypothesis 1, there was a decline in BPD from pre-to post-load sequence. In the current experiment, the decline in BPD did not depend on whether the load sequence was heavy or light, or whether the incentives were high and low, suggesting little influence of motivation and magnitude of fatigue on the BPD. Previously Hopstaken et al., (2015) report a steeper decline over the course of a visual 3-back task as compared to 2-back and 1-back tasks. This effect was accompanied by performance improvement in the 3-back task and performance declines in the 2-back and 1-back tasks. The steeper decline in BPD in the 3-back task was therefore attributed to learning effects. Similarly, Ayasse & Wingfield (2020) report a decline in BPD over the course of a SIN task that was steeper in individuals with poorer hearing thresholds. Inconsistent with the fatigue interpretation, there was an improvement in performance. The authors concluded that the BPD reflects a combination of different factors. In the current experiment, there was on average no change in performance from pre-to post-load sequence, independently of whether the load sequence was heavy or light. Self-reported tendency to quit listening did not change from the pre-to the post-load sequence trials either. This suggests that there is no evidence of any subjective experience of fatigue. On the other hand, self-report scale of tendency to quit may have been insensitive to capture the experience of fatigue due to a response bias. That is, participants may have been unwilling to admit their tendency to give up as part of social politeness (Grimm, 2010). Although it is difficult to completely rule out or pinpoint the contributions of fatigue, habituation, or learning to the overall decrease in BPD (arousal) that developed over the experiment, a decline in baseline is consistent with the decline of capacity, which is viewed as a hallmark of fatigue (Kahneman, 1973). Future studies could employ different (e.g. implicit) subjective measures to better assess the development of the subjective experience of fatigue.

Supporting Hypothesis 2, the average decline in PPD was larger after the heavy load sequence as compared to that after the light load sequence. One interpretation of this finding is that heavier load sequence results in larger mental fatigue, which diminishes the perceived value of investing effort in a greater extent than that in the light load sequence condition. Therefore, individuals subsequently invest less effort in listening. Previously Hornsby (2013) showed greater increase in secondary task RTs across a SIN task with concurrent memory load when participants were not wearing hearing aids as compared to when they were, suggesting that greater mental demands result in greater fatigue over time. Our results extend the findings of Hornsby (2013) with pupillometric measures, and support the understanding that reductions in mental demands of previous load can reduce fatigue effects. Another explanation for the larger decline in PPD after the heavy load sequence may be the learning of a listening strategy that is more economical on effort during the heavy load sequence condition. That is, to manage concurrent listening and memory demands, participants may have adopted an approach in listening that requires minimum effort and have continued with it throughout the post-load sequence blocks.

Supporting Hypothesis 3, on average, the decline in PPD from pre-to post-load sequence was larger when the incentives were low. This result suggests that motivational factors play a role in determining how much listeners exert effort after a period of effortful exertion. The moderating effect of monetary incentives can be interpreted in consideration of the FUEL and complementary models of motivational fatigue (Hockey, 2010; Müller & Apps, 2019; Pichora-Fuller et al., 2016). That is, after performing the load sequence, investing effort may have been perceived as worthwhile on trials with high monetary incentives and less worthwhile on trials with lower monetary incentives.

Overall, we did not observe any interaction between load sequence magnitude and monetary incentives on the PPD (Hypothesis 4 not supported); however, post-hoc analyses showed that the change in PPD from pre-to post-load sequence was only significant in the heavy-load-low-incentive condition. This result is in line with the understanding that the effect of fatigue is moderated by motivational factors (Hockey, 2010; Müller & Apps, 2019; Pichora-Fuller et al., 2016). Although learning effects on the decline in PPD from pre-to post-load sequence in the heavy load sequence condition should not be ruled out, they are insufficient to explain the effect of monetary incentives on the change in PPD. The current finding suggests that in a SIN task, fatigue effects are only visible when the willingness to perform well is low. In other words, the effect of fatigue on the PPD may be masked by motivational factors.

Whereas Wang et al (2018) have shown correlational evidence for the relationship between daily-life fatigue and PPD, the current results are the first to refine this understanding by demonstrating the impact of task-induced fatigue on PPD.

### Limitations

The study has some limitations that should be mentioned. Technical difficulties in the collection of pupil data led to the exclusion of 12 participants from the study, potentially limiting the power of our statistical analyses. The self-report measures that were used in the study may have not been adequate to capture all dimensions and components of motivation and fatigue, such as sensitivity to monetary rewards, or sleepiness and tiredness. Better such measures would have been useful in evaluating the extent to which the offered monetary incentives were effective in manipulating motivation, and to the nature of the mental state that the load sequence task has elicited. In this study the effects of motivation and task-induced fatigue were investigated in a sample of NH adults. The generalizability of the mechanisms explored here to listening-related daily-life fatigue in adults with HI is left unclear. Longitudinal studies utilizing physiological methods would be required if one wished to shed light on how task-induced fatigue relates to the development of daily-life fatigue in adults with HI.

## Conclusions

The present study demonstrates that task-induced fatigue and motivation interactively affect the capacity mobilized for listening. The BPD declined from pre-to post-load sequence, suggesting an overall decrease in arousal, but the decline did not scale with the magnitude of the previous mental load. The PPD declined from pre-to post-load sequence significantly only in the heavy load sequence-low monetary incentive condition. This supports the understanding that the investment of listening effort depends on an individual’s listening goals (Pichora-Fuller et al., 2016). The current results show that the influence of fatigue on effort is not uniform across different motivational states, suggesting that models of listening effort and listening-related fatigue may need to consider motivation (the willingness to invest effort) and (listening-related) fatigue as different parameters that can have a range of values.

## Data Availability

All data produced in the present study are available upon reasonable request to the authors

## Acknowledgements

We are grateful to Patrycja Książek, Renskje K. Hietkamp, Antoine Favre-Félix, Sara Al-ward, Eva Mathiesen, Julia Yevchenko, and Franziska Behnen for discussions, assistance in data-collection,. and technical experimental set-up.

This study has received funding from the European Union’s Horizon 2020 research and innovation programme under the Marie-Sklodowska-Curie grant agreement No 765329; the Medical Research Council [grant number MR/S003576/1]; and the Chief Scientist Office of the Scottish Government. There are no conflicts of interest, financial, or otherwise.

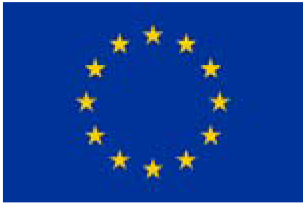

